## Supplementary Information for "The risk of SARS-CoV-2 outbreaks in low prevalence settings following the removal of travel restrictions"

**AUTHORS**

R. Sachak-Patwa[1], H.M. Byrne[1], L. Dyson[2,3], R.N. Thompson[2,3,*]

**AFFILIATIONS**

[1]Mathematical Institute, University of Oxford, Oxford, UK

[2]Mathematics Institute, University of Warwick, Coventry, UK

[3]Zeeman Institute for Systems Biology and Infectious Disease Epidemiology Research, University of Warwick, Coventry, UK

### <u>Outbreak risk metrics</u>

Here, we provide additional details about the metrics used in the main text for assessing the risk of an outbreak resulting from the introduction of an infectious host into the population.

*Instantaneous Outbreak Risk (IOR)*

The IOR represents the risk that an imported case initiates an outbreak as opposed to the virus simply fading out, under the assumption that pathogen transmissibility is fixed at its value when the imported case enters the population. If the introduction occurs at time $t$, when the infection rate is $\beta(1-\Lambda(t))$ and the time-varying reproduction number is $R_V(t)=\frac{\beta(1-\Lambda(t))}{\mu}$, then the outbreak probability can be derived by first denoting by $q_{I(t)}$ the probability that an outbreak does not occur starting from $I(t)$ infected individuals. Conditioning on whether the first event is an infection or a removal event gives

$$q_{I(t)}=\frac{\beta\big(1-\Lambda(t)\big)}{\beta\big(1-\Lambda(t)\big)+\mu}q_{I(t)+1}+\frac{\mu}{\beta\big(1-\Lambda(t)\big)+\mu}q_{I(t)-1}.$$

In particular, starting from a single introduction at time $t$,

$$q_1=\frac{\beta\big(1-\Lambda(t)\big)}{\beta\big(1-\Lambda(t)\big)+\mu}q_2+\frac{\mu}{\beta\big(1-\Lambda(t)\big)+\mu}q_0.$$

Since infections occur according to a branching process, $q_2={q_1}^2$. Furthermore, an outbreak will not occur if there are no infected individuals (i.e. $q_0=1$), so that

$$\beta\big(1-\Lambda(t)\big){q_1}^2-\Big(\beta\big(1-\Lambda(t)\big)+\mu\Big)q_1+\mu=0.$$

Solving this quadratic equation, and taking the minimal non-negative solution (as dictated by Markov chain theory when calculating the probability that a branching process fades out [1]) leads to the IOR,

$$\mathrm{IOR} = 1 - q_1 = \begin{cases} 0 \ \text{ for } R_V(t) \leq 1, \\ 1 - \dfrac{1}{R_V(t)} \ \text{ for } R_V(t) > 1. \end{cases} \tag{S1}$$

*Case Outbreak Risk (COR)*

Unlike the IOR, the COR accounts for changes in $\beta\big(1 - \Lambda(t)\big)$ that occur after the virus is introduced into the host population when assessing the outbreak risk. Similarly to above, the probability that an outbreak does not occur starting from $I$ infected individuals in the population at time $t$ is denoted by $q_I(t)$. Then, starting from a single introduction into the population at time $t$ and conditioning on the possible events events in the next $\Delta t$ days (where $\Delta t$ represents a short time period, so that at most a single event can occur in the interval $[t, t + \Delta t)$) gives

$$q_1(t) = \beta\big(1 - \Lambda(t)\big)\Delta t\, q_2(t + \Delta t) + \mu\Delta t\, q_0(t + \Delta t) + \big(1 - \beta\big(1 - \Lambda(t)\big)\Delta t - \mu\Delta t\big)q_1(t + \Delta t).$$

Since infections occur according to a branching process, $q_2(t + \Delta t) = q_1(t + \Delta t)^2$. Furthermore, an outbreak will not occur if there are no infected individuals (i.e. $q_0(t + \Delta t) = 1$), so that

$$q_1(t) = \beta\big(1 - \Lambda(t)\big)\Delta t\, q_1(t + \Delta t)^2 + \mu\Delta t\ + \big(1 - \beta\big(1 - \Lambda(t)\big)\Delta t - \mu\Delta t\big)q_1(t + \Delta t).$$

Rearranging this expression, and taking the limit $\Delta t \to 0$, gives equation (3) in the main text,

$$\frac{dq(t)}{dt} = \beta\big(1 - \Lambda(t)\big)q(t)\big(1 - q(t)\big) + \mu\big(q(t) - 1\big), \qquad \text{(S2)}$$

in which the variable $q_1(t)$ has been replaced by $q(t)$ for notational convenience. This equation can be solved numerically, after which the COR at time $t$ (i.e. the probability that an outbreak occurs) is given by $1 - q(t)$.

Solving equation (S2) numerically requires the value of $q(t)$ to be known at a single timepoint. In scenarios in which $R_V(t)$ is above one when the vaccine programme is completed, we solve equation (S2) backwards in time starting from the final condition $q(t^*) = \frac{1}{R_V(t^*)}$, where $t^*$ is the time at which the vaccination programme ends. The rationale for this choice is that the COR then matches the IOR for scenarios in which virus transmissibility does not change in future (e.g. the end of the vaccination programme). For scenarios in which $R_V(t)$ is instead less than one when the vaccination programme is completed, we set $q(0)$ so that $1 - q(0)$ matches the Numerical Outbreak Risk (NOR) at time $t = 0$ (see below), and then solve equation (S2) forwards in time starting from $t = 0$.

*Simulated Outbreak Risk (SOR)*

The SOR is calculated by simulating the stochastic branching process model $10{,}000$ times starting from a single infected individual introduced into the system at time $t$. The SOR is then given by the proportion of simulations in which the number of individuals infected simultaneously reaches the threshold $M = 100$ as opposed to fading out (i.e. the number of infected hosts reaches 100 before hitting zero).

To simulate the branching process model, we follow the following steps:

1. Set the initial time $t$, and set $I(t) = 1$.
2. Calculate the time of the next event, $t + \tau$, using the expression

$$\int_t^{t+\tau} (\beta(1 - \Lambda(s)) + \mu) I(s) \mathrm{d}s = -\ln(r_1),$$

   where $r_1$ is a random number sampled from a uniform distribution on (0,1).
3. Determine whether the next event is an infection event or a removal event. To do this, sample a second random number ($r_2$) from a uniform distribution on (0,1). If

$$r_2 < \frac{\beta(1 - \Lambda(t+\tau))}{\beta(1 - \Lambda(t+\tau)) + \mu},$$

   then the next event is an infection event; set $I(t+\tau) = I(t) + 1$. If instead the inequality above is not satisfied, then the next event is a removal event; set $I(t+\tau) = I(t) - 1$.
4. Repeat steps 2-3 while the outbreak is still ongoing and the threshold value of $M = 100$ has not been hit (i.e. $I(t) > 0$ and $I(t) < M$).

*Numerical Outbreak Risk (NOR)*

The NOR is analogous to the SOR, but with the advantage that it can be calculated without performing model simulations. To calculate the NOR, we define the vector $p(t) = \left(p_0(t), \dots, p_{M-1}(t), p_M(t)\right)^{\mathsf{T}}$, where, for $i \leq M-1$, $p_i(t)$ represents the probability that $I(t) = i$ and that the number infected has not reached threshold $M$ by time $t$. The variable $p_M(t)$ is defined as the probability that $I(t)$ has reached threshold $M$ by time $t$. As with the SOR, we again use a value of $M = 100$ in our analyses. Then, the Kolmogorov forward equations [2] are $\frac{dp(t)}{dt} = Q(t)p(t)$, where

$$Q(t) = \begin{pmatrix} 0 & \mu & 0 & \cdots & \cdots & 0 \\ 0 & -(\beta(1-\Lambda(t)) + \mu) & 2\mu & \cdots & \cdots & 0 \\ 0 & \beta(1-\Lambda(t)) & -(2\beta(1-\Lambda(t)) + 2\mu) & \cdots & \cdots & 0 \\ 0 & 0 & 2\beta(1-\Lambda(t)) & \cdots & \cdots & 0 \\ \vdots & \vdots & \vdots & \ddots & \vdots & \vdots \\ 0 & 0 & 0 & \vdots & (M-1)\mu & 0 \\ 0 & 0 & 0 & \vdots & -((M-1)\beta(1-\Lambda(t)) + (M-1)\mu) & 0 \\ 0 & 0 & 0 & \vdots & (M-1)\beta(1-\Lambda(t)) & 0 \end{pmatrix}.$$

In this matrix, the state $I(t) = M$ is an absorbing state (analogous to stopping the simulations as soon as $I(t)$ reaches the threshold $M$ when calculating the SOR). The probability that that are at least $M$ infected individuals at some stage following the introduction of a single infected host into the population at time $t$ is then given by $\mathrm{NOR}(t) = \lim_{s\to\infty}(p_M(s))$, where $p(t) = (0,1,0 \dots ,0,0)^{\mathsf{T}}$. The NOR is therefore calculated by solving the Kolmogorov forward equations numerically until the system converges to an equilibrium state.

**Individual-level variation in SARS-CoV-2 transmission**

Under the simple model of SARS-CoV-2 transmission considered in the main text, infected individuals are infectious for an exponentially distributed period (with mean $1/\mu$ days) and generate new infections at rate $\beta(1 - \Lambda(t))$ per day. Using this model, we found that the risk of outbreaks is likely to be greater than zero in low prevalence settings when NPIs are removed, even following vaccination programmes in which significant numbers of individuals are fully vaccinated. Here, we test the robustness of this conclusion to variation in the assumed level of heterogeneity in transmission between different infected individuals.

In the analysis in the main text, when vaccination programmes are completed, the offspring distribution (the number of infections generated by each infected individual) follows a geometric distribution with mean $R_V(t) = \frac{\beta(1-\eta_2 v)}{\mu}$ (we denote this final value of $R_V(t) = \frac{\beta(1-\eta_2 v)}{\mu}$ by $R_V$ for the remainder of this section). Since virus transmissibility does not then change in the future, the IOR is appropriate for assessing the risk of outbreaks.

In reality, however, there is significant variation in the number of infections generated by different infected individuals. The potential that "superspreading" events occur leads to a longer tailed offspring distribution than that of a geometric distribution. To allow for this in epidemiological models, a negative binomial distribution can be used instead of a geometric distribution to characterise the offspring distribution.

Here, we explore the outbreak risk at the end of the vaccination programmes in the Isle of Man and Israel using a negative binomial offspring distribution. Specifically, the probability that an infected individual generates $x$ secondary cases is given by

$$P(X = x) = \frac{\Gamma(k + \mathrm{x})}{x!\,\Gamma(k)} \left(\frac{R_V}{R_V + k}\right)^{x} \left(1 + \frac{R_V}{k}\right)^{-k},$$

where $k$ is the dispersion parameter [3]. When $k = 1$, this is equivalent to the geometric distribution assumed in the main text, but when $k < 1$ then there is more individual-level variation in transmission (i.e. a higher potential for superspreading).

Under this more complex transmission model, the probability that an infected individual, introduced into the population at any time following the vaccination programme, initiates an outbreak driven by sustained local transmission, is given by

$$\mathrm{IOR} = 1 - q,$$

where $q$ is the minimal non-negative solution of

$$q = \left(1 + \frac{R_V(1-q)}{k}\right)^{-k},$$

as described by Lloyd-Smith *et al.* [3] and Nishiura *et al.* [4]. As noted above, when the dispersion parameter $k = 1$, then the offspring distribution is geometric, in which case the IOR is given by the expression in equation (S1) (with $R_V(t) = R_V$, since the vaccination programme is assumed to have been completed).

We estimated the IOR at the ends of vaccination campaigns in the Isle of Man and Israel for different values of the dispersion parameter, $k$ (Fig S4). As in the main text, we considered two distinct scenarios corresponding to different mean values of $R_0$ (these values are $R_0 = 3$ and $R_0 = 5$). Blue dots in Fig S4 correspond to $k = 1$ (so that the IOR there matches the IOR values at the ends of the vaccination programmes in the corresponding panels of Fig 3 in the main text), and red dots in Fig S4 correspond to $k = 0.1$ (which is consistent with some published estimates of $k$ for SARS-CoV-2 transmission [5]).

In each scenario, once vaccination programmes have been completed, the IOR is greater than zero for realistic values of the dispersion parameter, $k$. The IOR is particularly large in the scenarios for which the mean value of $R_0 = 5$. We expect these scenarios to reflect the current and future risks most accurately, due to the emergence of variants that are more transmissible than the original SARS-CoV-2 virus. The results in Fig S4 support our main conclusion that the risk of outbreaks in low prevalence settings (once NPIs are removed) is unlikely to be eliminated by vaccination. Surveillance strategies that aim to identify infected inbound travellers in locations with low infection prevalence are of clear importance, even once vaccination programmes are completed.

### Supplementary figures

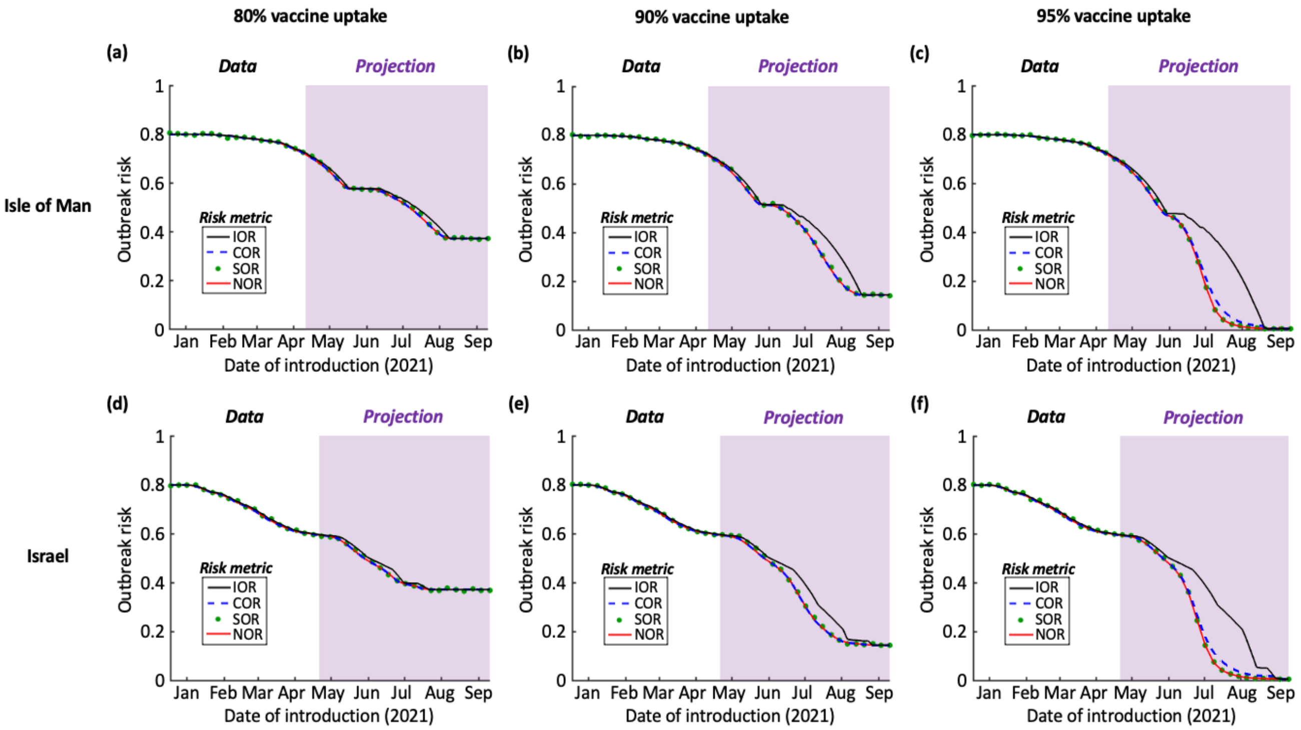


Figure S1. The effect of the assumed vaccine uptake on the outbreak risk. (a) Isle of Man. Same result as Fig 3b in the main text (vaccine uptake $\nu = 0.8$). (b) Analogous panel to Fig 3b in the main text, but with vaccine uptake $\nu = 0.9$. (c) Analogous panel to Fig 3b in the main text, but with vaccine uptake $\nu = 0.95$. (d) Israel. Analogous panel to Fig 3d in the main text, but with vaccine uptake $\nu = 0.8$. (e) Analogous panel to Fig 3d in the main text, but with vaccine uptake $\nu = 0.9$. (f) Analogous panel to Fig 3d in the main text, but with vaccine uptake $\nu = 0.95$. Ticks on the x-axes refer to the starts of the months labelled.

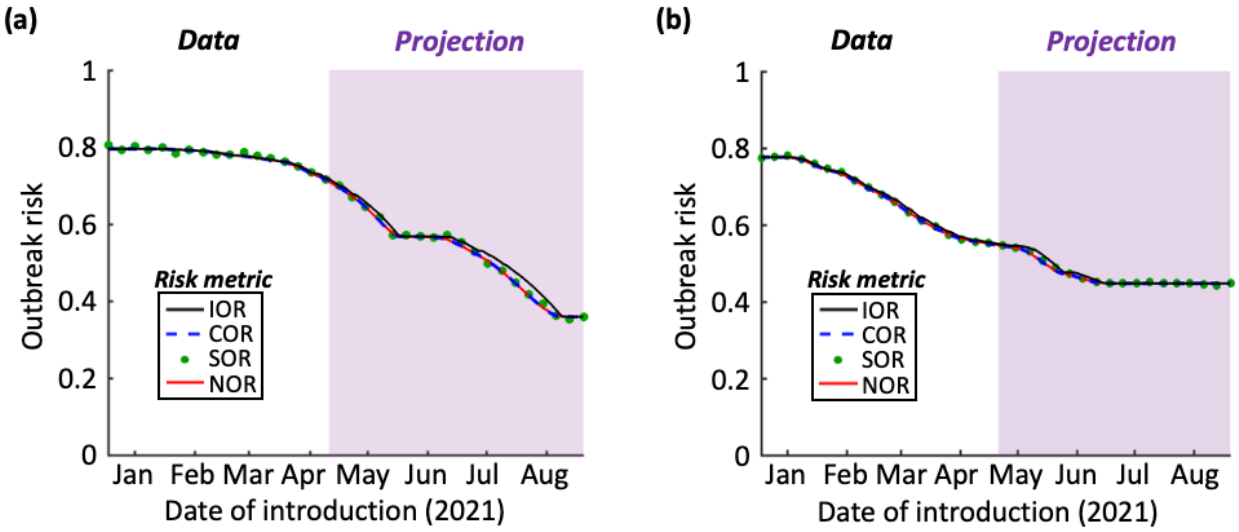


Figure S2. The effect of immunity due to prior infections on the outbreak risk. (a) Isle of Man. Analogous panel to Fig 3b in the main text, but with $R_0$ reduced by 1.4% (an approximation of the proportion of the population who had been infected before 1st May 2021, based on 1,154 confirmed cases within a population of $N = 84{,}500$). (b) Israel. Analogous panel to Fig 3d in the main text, but with $R_0$ reduced by 9.6% (an approximation of the proportion of the population who had been infected before 1st May 2021, based on 838,000 confirmed cases within a population of $N = 8{,}772{,}800$). Ticks on the x-axes refer to the starts of the months labelled.

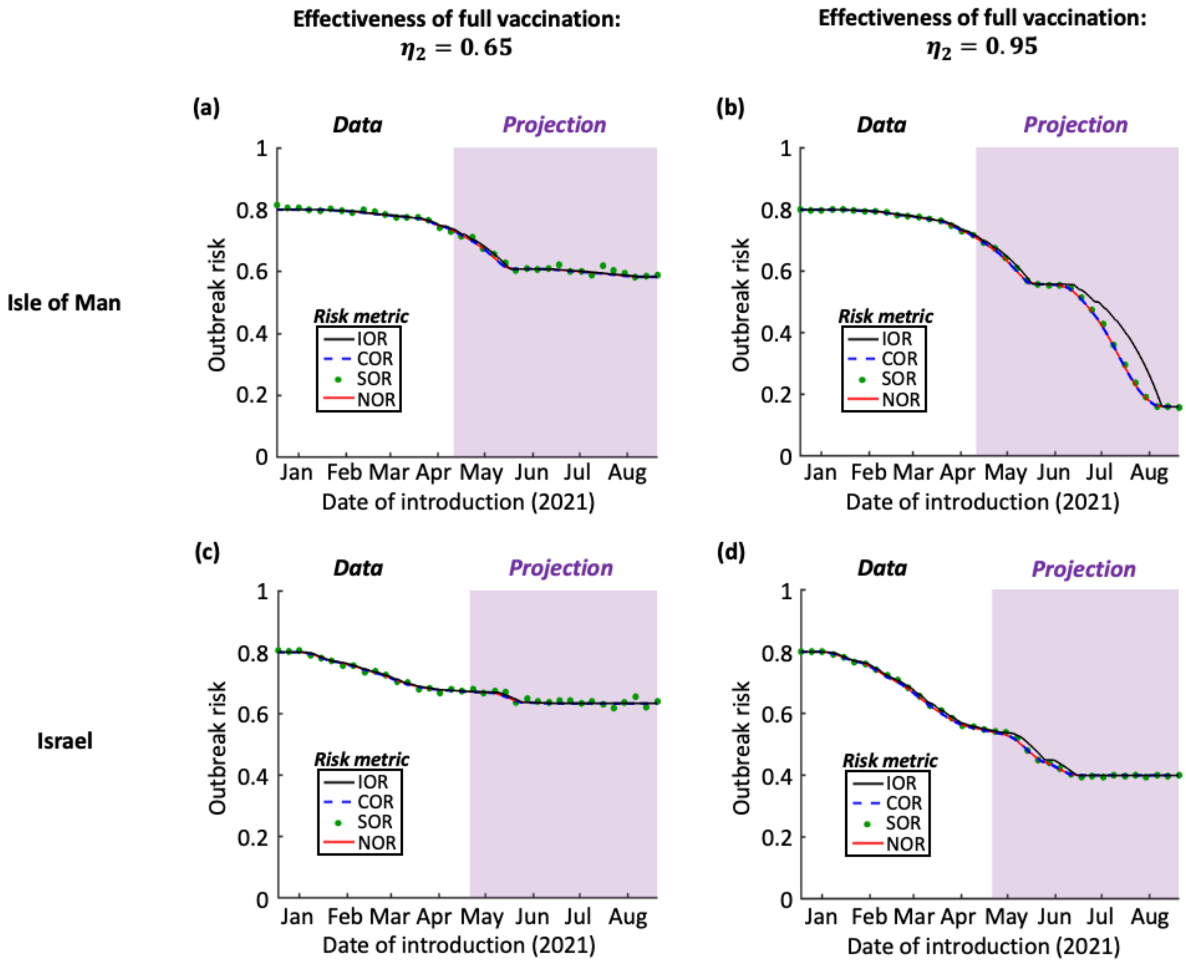


Figure S3. The effect of the assumed effectiveness of two vaccine doses on the outbreak risk. (a) Isle of Man. Analogous panel to Fig 3b in the main text, but with $\eta_2 = 0.65$. (b) Analogous panel to Fig 3b in the main text, but with $\eta_2 = 0.95$. (c) Israel. Analogous panel to Fig 3d in the main text, but with $\eta_2 = 0.65$. (d) Analogous panel to Fig 3d in the main text, but with $\eta_2 = 0.95$. Ticks on the x-axes refer to the starts of the months labelled.

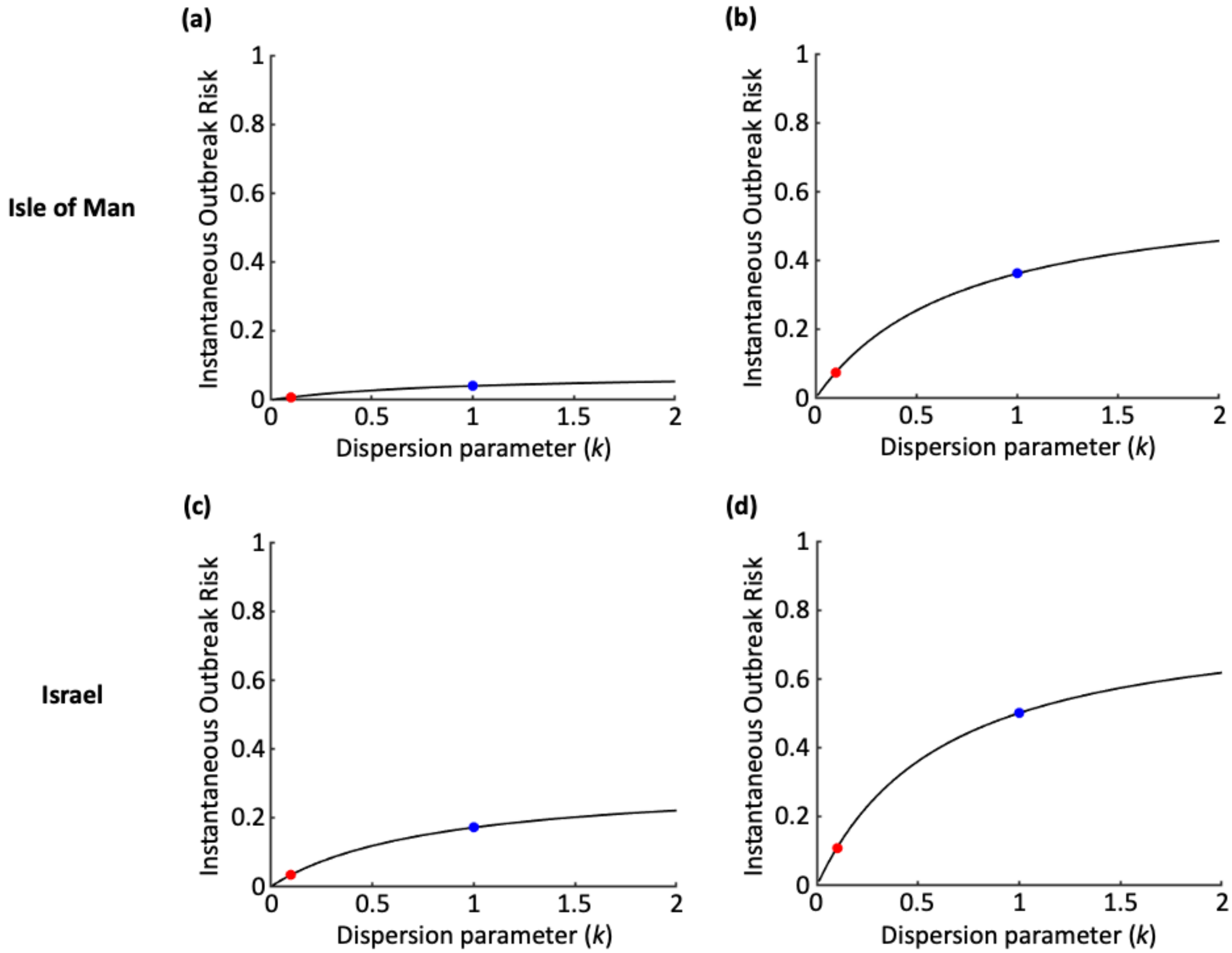


Figure S4. The effect of individual-level variation in transmission between different infected individuals (determined by the dispersion parameter, $k$, of the negative binomial offspring distribution) on the IOR following the completion of the vaccination programmes. (a) Isle of Man, with mean $R_0 = 3$ at the beginning of the vaccination programme (as in Fig 3a of the main text). (b) Isle of Man, with mean $R_0 = 5$ at the beginning of the vaccination programme (as in Fig 3b of the main text). (c) Israel, with mean $R_0 = 3$ at the beginning of the vaccination programme (as in Fig 3c of the main text). (d) Israel, with mean $R_0 = 5$ at the beginning of the vaccination programme (as in Fig 3d of the main text). Blue dots correspond to $k = 1$ (the underlying assumption of the simple branching process model used in the main text) and red dots correspond to $k = 0.1$ (as estimated for SARS-CoV-2 transmission in [5], although larger values of $k$ have been estimated in some studies [6,7]). The values of other parameters are given in Table 1 of the main text.
